## Supplementary Tables for "Epidemiology of Atrial Fibrillation in the *All of Us* Research Program"

**SUPPLEMENTARY MATERIALS**

Supplementary Table S1. Concept IDs used to identify atrial fibrillation in the electronic health records, *All of Us* Research Program, 2017-2019

| **Description** | **OMOP Concept ID** | **SNOMED Code** |
| --- | --- | --- |
| Atrial fibrillation | 313217 | 49436004 |
| Atrial fibrillation and flutter | 4108832 | 195080001 |
| Atrial fibrillation with rapid ventricular response | 44782442 | 120041000119109 |
| Chronic atrial fibrillation | 4141360 | 426749004 |
| Controlled atrial fibrillation | 4117112 | 300996004 |
| ECG: atrial fibrillation | 4064452 | 164889003 |
| Lone atrial fibrillation | 4119601 | 233910005 |
| Paroxysmal atrial fibrillation | 4154290 | 282825002 |
| Permanent atrial fibrillation | 4232691 | 440028005 |
| Persistent atrial fibrillation | 4232697 | 440059007 |
| Rapid atrial fibrillation | 4199501 | 314208002 |

OMOP: Observational Medical Outcomes Partnership; SNOMED: Systematized Nomenclature of Medicine.

Supplementary Table S2. Participant characteristics by study component participation. Values correspond to mean (standard deviation) or N (%), *All of Us* Research Program 2017-2019

|  | Overall | Both EHR and survey data | Only EHR data | Only survey data | No EHR, no survey data |
| --- | --- | --- | --- | --- | --- |
| N | 173,101 | 20,683 | 71,636 | 14,800 | 65,982 |
| Age, years | 52 (17) | 55 (16) | 52 (17) | 53 (17) | 51 (17) |
| Female sex | 105,524 (61%) | 13,939 (67%) | 43,500 (61%) | 9,387 (63%) | 38,698 (59%) |
| Race/ethnicity |  |  |  |  |  |
| Non-Hispanic White | 116,241 (67%) | 18,024 (87%) | 43,920 (61%) | 13,182 (89%) | 41,115 (62%) |
| Non-Hispanic Black | 44,993 (26%) | 1,559 (8%) | 22,876 (32%) | 680 (5%) | 19,878 (30%) |
| Non-Hispanic Asian | 7,338 (4%) | 696 (3%) | 2,748 (4%) | 599 (4%) | 3,295 (5%) |
| Hispanic | 4,529 (3%) | 404 (2%) | 2,092 (3%) | 339 (2%) | 1,694 (3%) |
| BMI, kg/m^2^ | 29.0 (6.7) | 28.3 (6.3) | 29.4 (6.8) | 27.9 (6.1) | 29.0 (6.7) |
| SBP, mmHg | 128 (19) | 127 (17) | 128 (19) | 127 (17) | 129 (19) |
| DBP, mmHg | 78 (12) | 77 (11) | 79 (12) | 77 (11) | 79 (12) |
| Ever smoker | 70,320 (41%) | 7,098 (34%) | 32,621 (46%) | 4,690 (32%) | 25,911 (39%) |
| Diabetes | 15,587 (9%) | 2,527 (12%) | 13,060 (18%) | -- | -- |
| Heart failure | 5,177 (3%) | 721 (3%) | 4,456 (6%) | -- | -- |
| CHD | 8,912 (5%) | 1,584 (8%) | 7,328 (10%) | -- | -- |
| Stroke | 490 (0.3%) | 81 (0.4%) | 409 (0.6%) | -- | -- |

BMI: body mass index; CHD: coronary heart disease; DBP: diastolic blood pressure; SBP: systolic blood pressure.

Supplementary Table S3. Association of age, sex, race/ethnicity and cardiovascular risk factors with prevalent atrial fibrillation among *All of Us* participants with EHR and medical history survey data (N = 20,683), *All of Us* Research Program, 2017-2019

|  | Odds ratio (95% CI) |
| --- | --- |
|  | Model 1* |
| Age group |  |
| <40 | 1 (Ref.) |
| 40-49 | 2 (1.4, 3.0) |
| 50-59 | 3.4 (2.5, 4.7) |
| 60-69 | 7.7 (5.8, 10.2) |
| 70-79 | 11.9 (8.9, 15.9) |
| 80+ | 24.1 (17.4, 33.5) |
| Sex |  |
| Female | 1 (Ref.) |
| Male | 1.8 (1.6, 2.0) |
| Race / ethnicity |  |
| Hispanic | 0.91 (0.51, 1.6) |
| NH Asian | 0.55 (0.34, 0.89) |
| NH Black | 0.62 (0.46, 0.82) |
| NH White | 1 (Ref.) |
|  | Model 2** |
| BMI, per 5 kg/m^2^ | 1.1 (1.1, 1.2) |
| SBP, per 20 mmHg | 0.88 (0.80, 0.96) |
| DBP, per 10 mmHg | 1.1 (1.0, 1.1) |
| Ever smoking | 1.1 (0.97, 1.2) |
| Diabetes | 0.94 (0.80, 1.1) |
| Heart failure | 5.1 (4.2, 6.2) |
| Coronary heart disease | 2.2 (1.9, 2.6) |
| Stroke | 2.6 (1.4, 4.8) |

* Logistic regression including age (categories), sex, and race/ethnicity. ** Logistic regression including age (continuous), sex, race/ethnicity and all other covariates in the table.

Supplementary Table S4. Age, sex, and race/ethnicity prevalence of AF in the *All of Us* Research Program and selected epidemiologic studies in the United States. Values correspond to prevalence per 100 persons.

| *All of Us* Research Program | | | ATRIA^1^ | | | Medicare 2007^2^ | |
| --- | --- | --- | --- | --- | --- | --- | --- |
|  | Survey | EHR |  | Females | Males |  | |
| Age |  |  | Age |  |  | Age |  |
| <40 | 0.9 | 0.3 | <55 | 0.1 | 0.2 |  |  |
| 40-49 | 2.2 | 1.3 | 55-59 | 0.4 | 0.9 |  |  |
| 50-59 | 3.7 | 2.4 | 60-64 | 1.0 | 1.7 |  |  |
| 60-69 | 7.5 | 5.5 | 65-69 | 1.7 | 3.0 | 66-69 | 3.1 |
| 70-79 | 13.0 | 11.1 | 70-74 | 3.4 | 5.0 | 70-74 | 5.8 |
| 80-89 | 22.0 | 19.1 | 75-79 | 5.0 | 7.3 | 75-79 | 9.4 |
|  |  |  | 80-84 | 7.2 | 10.3 | 80-84 | 13.1 |
|  |  |  | ≥85 | 9.1 | 11.1 | 85-89 | 16.3 |
|  |  |  |  |  |  | ≥90 | 17.5 |
| Sex |  |  | Sex |  |  | Sex |  |
| Female | 4.3 | 3.0 | Female | 1.1 | | Female | 7.4 |
| Male | 8.8 | 6.1 | Male | 0.8 | | Male | 10.4 |
| Race/  ethnicity |  |  | Race/  ethnicity |  |  | Race/  ethnicity |  |
| Hispanic | <2.7 | 1.8 |  |  |  |  |  |
| NH Asian | 2.1 | 1.8 |  |  |  |  |  |
| NH Black | 2.8 | 2.2 | Black | 1.5 | | Black | 4.6 |
| NH White | 6.3 | 5.2 | White | 2.2 | | White | 9.1 |

ATRIA: AnTicoagulation and Risk Factors In Atrial Fibrillation. NH: Non-Hispanic

Supplementary Table S5. Age, sex, and race/ethnicity incidence rates of AF in the *All of Us* Research Program and selected epidemiologic studies in the United States. Values correspond to incidence rate per 1000 person-years.

| *All of Us*  Research Program | | CHS^3^ | | | FHS^4^ | | | ARIC study^5^ | | | Medicare 2007^2^ | |
| --- | --- | --- | --- | --- | --- | --- | --- | --- | --- | --- | --- | --- |
|  |  |  | Females | Males |  | Females | Males |  | | |  | |
| Age |  | Age |  |  | Age |  |  | Age |  | Age | |  |
| <40 | <0.6 |  |  |  |  |  |  |  |  |  | |  |
| 40-49 | <1.2 |  |  |  |  |  |  | 45-49 | 0.6 |  | |  |
| 50-59 | 2.2 |  |  |  | 55-64 | 5 | 7 | 50-59 | 1.8 |  | |  |
| 60-69 | 4.1 | 65-69 | 10.9 | 12.3 | 65-74 | 10 | 18 | 60-69 | 5.3 | 66-69 | | 12.9 |
| 70-79 | 7.6 | 70-74 | 9.1 | 22.8 | 75-84 | 30 | 35 | 70-89 | 12.2 | 70-74 | | 18.8 |
| 80-89 | 12.3 | 75-79 | 23.1 | 34.8 | 85-94 | 65 | 77 | ≥80 | 38.7 | 75-79 | | 28.8 |
|  |  | ≥80 | 25.1 | 58.7 |  |  |  |  |  | 80-84 | | 38.3 |
|  |  |  |  |  |  |  |  |  |  | 85-89 | | 53.5 |
|  |  |  |  |  |  |  |  |  |  | ≥90 | | 68.9 |
| Sex |  | Sex |  |  | Sex |  |  | Sex |  | Sex | |  |
| Female | 2.2 | Female | 14.1 | | Female | 11.3 | | Female | 3.7 | | Female | 24.7 |
| Male | 4.1 | Male | 26.4 | | Male | 14.5 | | Male | 6.1 | | Male | 33.9 |
| Race/  ethnicity |  | Race/  ethnicity |  |  |  |  |  | Race/  ethnicity |  | Race/  ethnicity | |  |
| Hispanic | <5.9 |  |  |  |  |  |  |  |  |  | |  |
| NH Asian | <4.1 |  |  |  |  |  |  |  |  |  | |  |
| NH Black | 1.5 | Black | 12.0 | |  |  |  | Black | 3.3 | Black | | 22.1 |
| NH White | 3.6 | Non-Black | 19.5 | |  |  |  | White | 5.2 | White | | 29.4 |

ARIC: Atherosclerosis Risk in Communities. CHS: Cardiovascular Health Study. FHS: Framingham Heart Study. NH: Non-Hispanic.
